# Evaluation of Convalescent Plasma Versus Standard of Care for the Treatment of COVID-19 in Hospitalized Patients: study protocol for a phase 2 randomized, open-label, controlled, multicenter trial

**DOI:** 10.1101/2020.07.31.20165720

**Authors:** Elena Diago-Sempere, José Luis Bueno, Aránzazu Sancho-López, Elena Múñez Rubio, Ferrán Torres, Rosa Malo de Molina, Ana Fernández-Cruz (AFC), Isabel Salcedo de Diego, Ana Velasco-Iglesias, Concepción Payares-Herrera, Inmaculada Casas Flecha, Cristina Avendaño-Solà, Rafael Duarte Palomino, Antonio Ramos-Martínez, Belén Ruiz-Antorán

## Abstract

**Background:** COVID-19 is a respiratory disease caused by a novel coronavirus (SARS-CoV-2) and causes substantial morbidity and mortality. At the time this clinical trial was planned, there were no available vaccine or therapeutic agents with proven efficacy, but the severity of the condition prompted the use of several pharmacological and non-pharmacological interventions.

It has long been hypothesized that the use of convalescent plasma (CP) from infected patients who have developed an effective immune response is likely to be an option for the treatment of patients with a variety of severe acute respiratory infections (SARI) of viral etiology. The aim of this study is to assess the efficacy and safety of convalescent plasma in adult patients with severe COVID-19 pneumonia.

**Methods/Design:** The ConPlas-19 study is a multicenter, randomized, open-label controlled trial. The protocol has been prepared in accordance with the SPIRIT (Standard Protocol Items: Recommendations for Interventional Trials) guidelines. The study has been planned to include 278 adult patients hospitalized with severe COVID-19 infection not requiring mechanical ventilation (invasive or non-invasive). Subjects are randomly assigned in a 1:1 ratio (139 per treatment arm), stratified by center, to receive intravenously administered CP (single infusion) plus SOC or SOC alone, and are to be followed for 30 days. The primary endpoint of the study is the proportion of patients that progress to categories 5, 6 or 7 (on the 7-point ordinal scale proposed by the WHO) at day 15. Interim analyses for efficacy and/or futility will be conducted once 20%, 40%, and 60% of the planned sample size are enrolled and complete D15 assessment.

**Discussion:** This clinical trial is designed to evaluate the efficacy and safety of passive immunotherapy with convalescent plasma for the treatment of adult patients hospitalized with COVID-19. The results of this study are expected to contribute to establishing the potential place of CP in the therapeutics for a new viral disease.

**Trial registration:** Trial registration at clinicaltrials.gov; Registration Number: NCT04345523; https://clinicaltrials.gov/ct2/show/NCT04345523; Registered on 30 March, 2020. First posted date: April 14, 2020.

## Background

COVID-19 is a respiratory disease caused by a novel coronavirus (SARS-CoV-2) and causes substantial morbidity and mortality. At the time the study was planned there were no vaccines to prevent COVID-19 or infection with SARS-CoV or therapeutic agent with demonstrated efficacy as specific treatment for COVID-19.

Convalescent plasma (CP) from infected patients who have developed an immune response is likely to be an option for the treatment of patients with a variety of severe viral diseases. This would include patients in the most recent epidemics with coronaviruses, SARS1 in 2003 and MERS in 2012, and potentially as well patients in the current COVID-19 pandemic. Despite suggesting safety and potential efficacy, the available evidence has the major limitation of being based on predominantly low-quality uncontrolled studies (1). Here we present a summary of the rationale and justification for conducting a multicenter, randomized clinical trial of CP therapy in COVID-19 hospitalized patients.

Passive immunotherapy involves the administration of antibodies against a given agent to a susceptible individual with the purpose of preventing or treating an infectious disease caused by that agent. Historically CP has been used in outbreaks of poliomyelitis, measles, mumps, influenza (1918 H1N1 and 2009-2010 H1N1),and 2013 Ebola (2). In addition, although less readily available and requiring more complex manufacturing than CP, conventional and hyperimmune immunoglobulins are used in clinical practice on a number of infections such as respiratory syncytial virus, hepatitis B and others(3).

Currently, the only source of antibodies available for immediate use against SARS-CoV-2 is human CP. This is a readily available resource during an epidemic crisis even in low-income countries, as it uses the infrastructure and means developed for blood transfusions. In addition, as more individuals contract COVID-19 and recover, the number of potential donors will continue to increase in all areas where COVID-19 epidemic is present(4).

The experience with severe acute respiratory infections (SARI) caused by a coronavirus is rather recent in a number of epidemics in the twenty first century. Human CP was used in patients from both SARS-1 in 2003 and MERS in 2012. Overall, the experience showed that CP is safe and likely to reduce mortality in patients with coronavirus-related SARI. The largest study with 80 patients with SARS in Hong-Kong in 2003 (5) and subsequent publications (6, 7), point out that earlier administration after symptom onset is more effective, particularly before day 14, prior to seroconversion in patients remaining PCR test positive.

From this background and rationale, we have developed this study with the objective of evaluating the safety and efficacy of CP in hospitalized adult patients with severe COVID-19. In the midst of a worldwide pandemic of SARS-CoV-2 and COVID-19, CP was hypothesized to represent a potential effective therapeutic option with a favorable safety profile for these patients.

### Objectives

The trial objective is to evaluate the clinical efficacy and safety of Convalescent Plasma combined with standard of care (SOC) compared with SOC alone in adult patients with severe COVID-19.

### Study design

This is a phase 2, parallel group, randomized, open-label, controlled, superiority, multicenter clinical trial.

The protocol has been prepared in accordance with the SPIRIT (Standard Protocol Items: Recommendations for Interventional Trials) guidelines (Figure 1).

**Figure 1.**
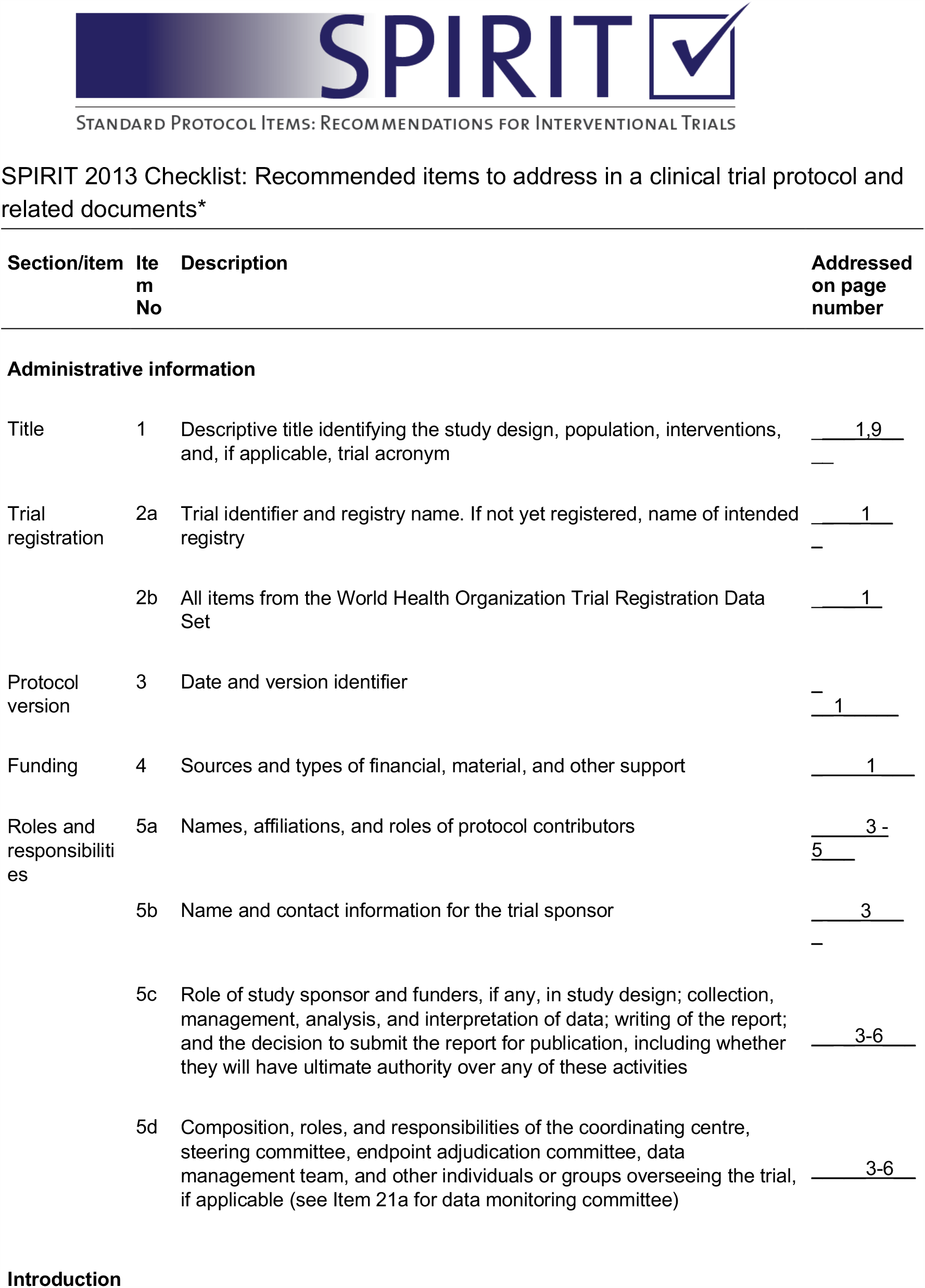
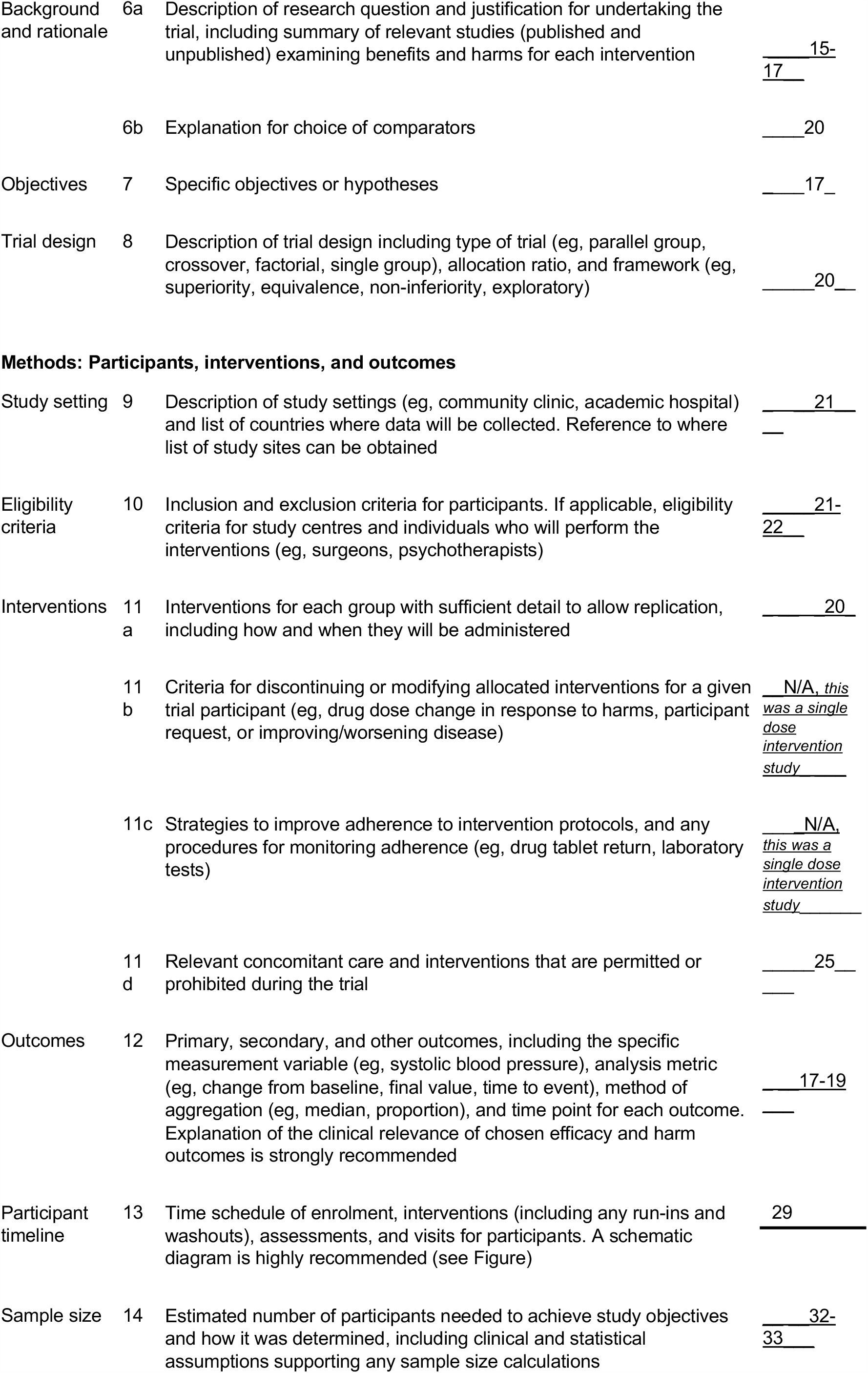
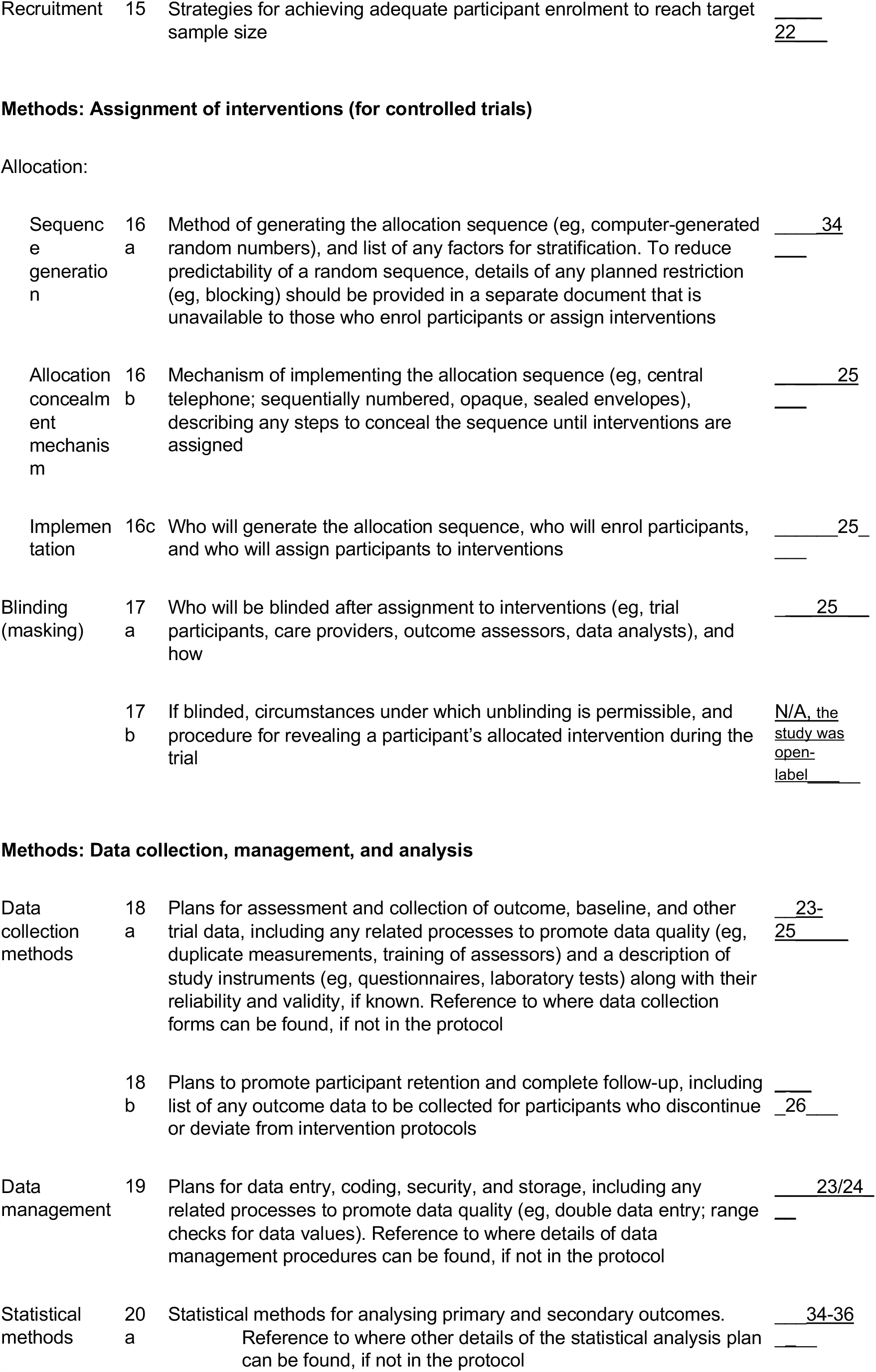
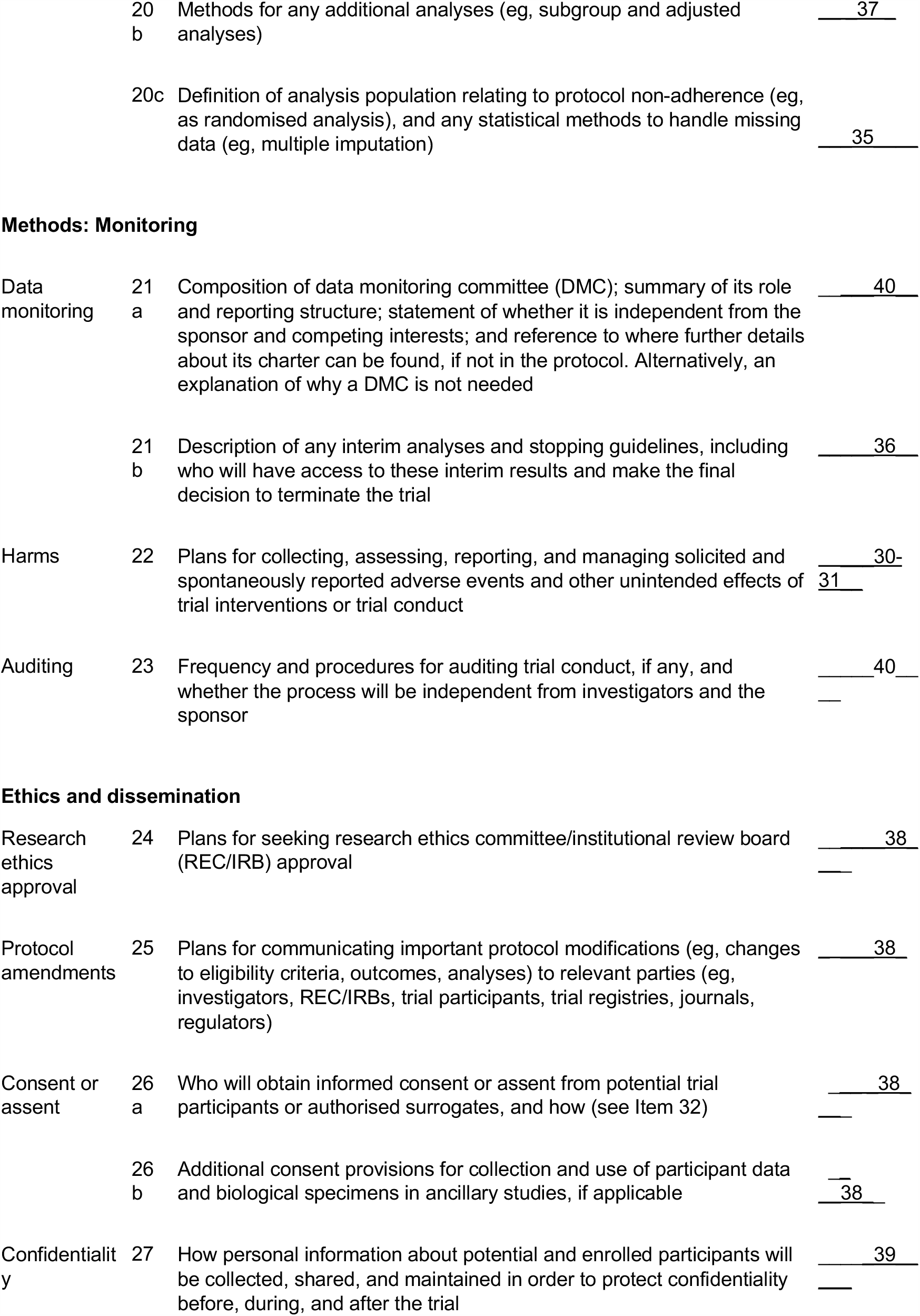
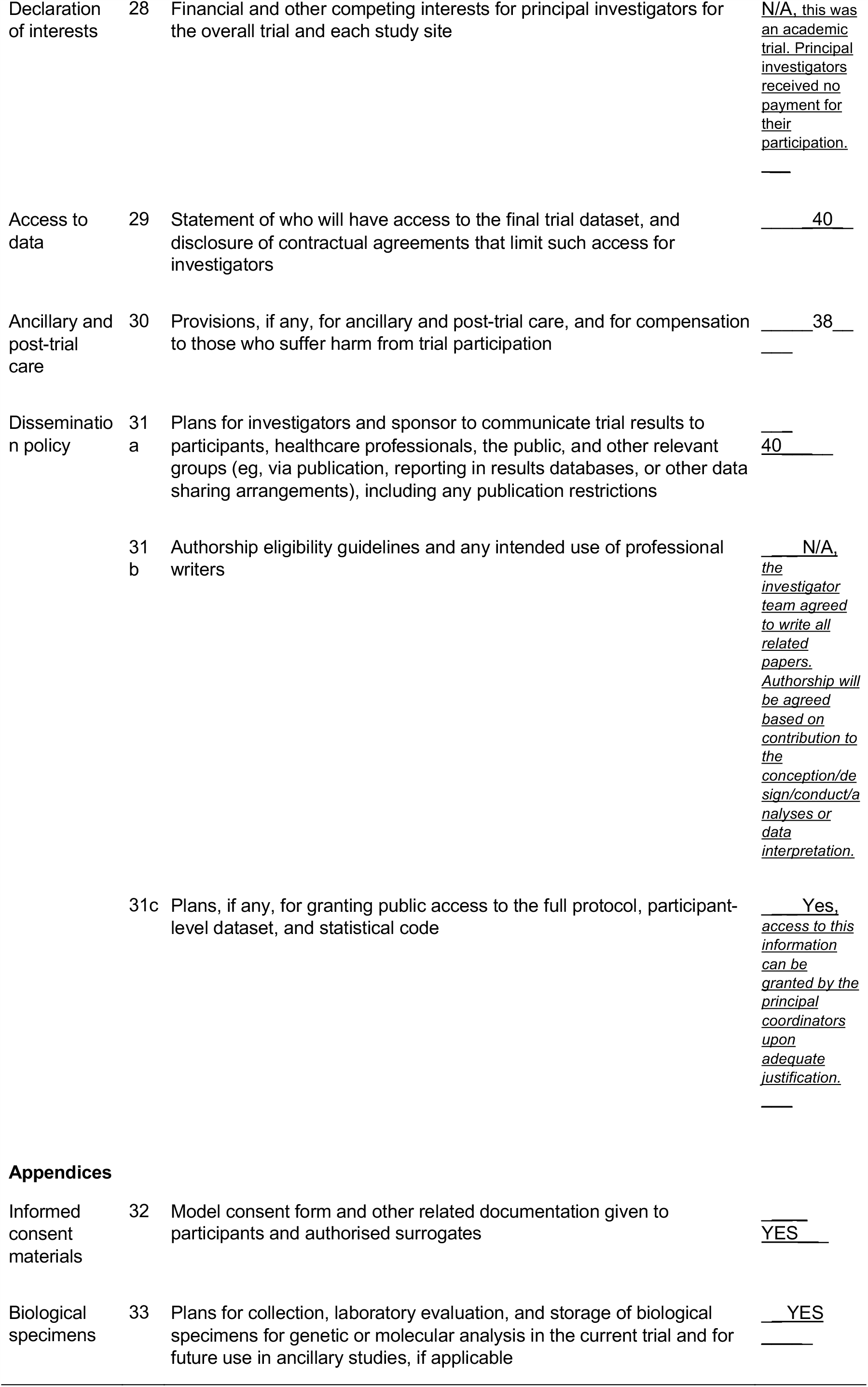
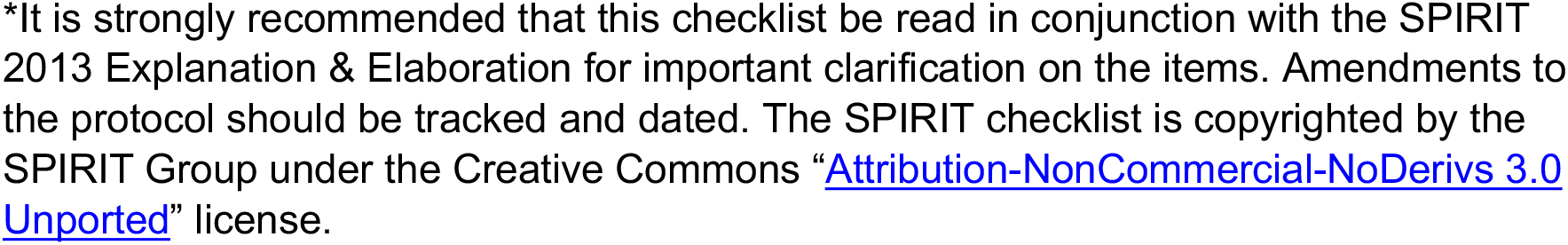

Approximately 278 patients (139 per arm) with severe SARS-CoV-2 pneumonia will be enrolled at 32 centers. Individuals fulfilling selection criteria will be randomized to receive CP (+SOC) or SOC alone at a ratio of 1:1. Also, approximately 140-200 CP donors will be recruited.

The study has been planned with a sequential design. Interim analyses for safety monitoring and for comprehensive efficacy/futility will be conducted when 20%, 40%, 60% and 80% of patients have been recruited and completed D15 primary endpoint assessment, or at the discretionary criteria of the Data Safety Monitoring Board (DSMB) when needed.

### Methods: participants, interventions, and outcomes

#### Participating centers

Study clinical sites included 32 hospitals across the different regions in Spain. ISCIII (Centro Nacional de Microbiología, Instituto de Salud Carlos III) centralized the antibodies and PCR microbiological testing from patients and donors. Transfusion services or centers at the hospitals or Autonomous Communities and National Army Transfusion Center are also essential participants in this trial.

### Eligibility criteria

#### Eligibility criteria for patients

*Inclusion criteria are:*

1. Written informed consent prior to performing study procedures. Witnessed oral consent will be accepted in order to avoid paper handling, which increased the risk of transmission of the infection. Written consent by patient or representatives will be obtained as soon as possible.
2. Male or female adult patient ≥ 18 years of age at the time of enrolment.
3. Laboratory-confirmed SARS-CoV-2 infection as determined by PCR in naso/oropharyngeal swabs or any other relevant specimen obtained in the ongoing COVID-19 symptomatic period.
4. Patients requiring hospitalization for COVID-19 without mechanical ventilation (invasive or non-invasive) or high flow oxygen devices and at least one of the following:
  - Radiographic evidence of pulmonary infiltrates by imaging (chest x-ray, CT scan, etc.) OR
  - Clinical assessment (evidence of rales/crackles on exam) and SpO2 ≤ 94% on room air that requires supplemental oxygen.
5. No more than 12 days between the onset of symptoms (fever or cough) and treatment administration day.

*Exclusion criteria are:*

1. Requiring mechanical ventilation (invasive or non-invasive) or high flow oxygen devices at screening.
2. More than 12 days since symptoms (fever or cough) onset.
3. Participation in any other clinical trial of an experimental treatment for COVID-19.
4. In the opinion of the clinical team, progression to death is imminent and inevitable within the next 24 hours, irrespective of the provision of treatments.
5. Any incompatibility or allergy to the administration of human plasma.
6. Stage 4 severe chronic kidney disease or requiring dialysis (i.e. eGFR<30).

#### Eligibility criteria for donors

*Inclusion criteria are:*

1. Subjects willing and able to provide written informed consent.
2. Fulfilling all the current requirements to be a plasma apheresis donor according to the regulations for donation of blood products (European Guidelines and RD 1088/2005 in Spain).
3. Absence of COVID-19 symptoms within the last 14 days.
4. Anti SARS-CoV-2 IgG antibodies detectable in peripheral blood.
5. ≥18 years of age at time of donation.
6. Weight > 50kg and good vein access are standard criteria, for which exceptions could be considered according to the criteria of the blood bank and hematologist.

*Exclusion criteria are:*

1. Plasmapheresis in the previous seven days.
2. Whole blood donation in the previous 30 days.
3. Donation of more than 25 liters of plasma in the previous 12 months.

#### Informed consent

Investigators will obtain the subject’s informed consent in accordance with Spanish Law 14/2007 on Biomedical Investigation and the internationally ethical accepted guidelines.

Patients will receive a concise and focused presentation of key information about the clinical trial, orally, and a written informed consent form will be handled to the patient. Due to paper handling limitation in COVID wards, oral witnessed consent will be accepted before entering into the trial, with written documentation in the patient clinical record. If possible, written consent form will be obtained from the patient himself or acceptable representatives,at a later time.

Donors will receive concise information about the clinical trial and will give written informed consent before donating convalescent plasma.

#### Additional consent provisions

The consent form includes provisions for research data and residual samples to be stored for future scientific research on COVID-19. These future studies will be previously evaluated by a Research Ethics Committee and will comply with the applicable ethical and legal requirements.

### Study interventions

#### Intervention description

All trial participants will receive SOC for COVID-19. The control arm is SOC for COVID-19. In the treatment arm, patients will also receive intravenous pathogen reduced CP from patients recovered from COVID-19 (designated as donors) as add-on therapy to SOC.

In the current status of a worldwide pandemic for which we have no approved vaccines or drugs, for the purpose of this trial SOC will include any medicinal products being used in clinical practice (e.g. lopinavir/ritonavir; darunavir/cobicistat; hydroxy/chloroquine, tocilizumab, remdesivir, allowed as SOC when its use outside clinical trials was permitted), other than those used as part of another clinical trial.

Donor assessment, pathogen reduced plasma collection and production will be performed by hospital Transfusion Services and Regional Transfusion Centres. Local organization will be adapted to the existing structure at the regional level.

#### Criteria for discontinuing or modifying allocated interventions

A patient may be removed from the study treatment for the reasons mentioned below, although whenever possible the patient should be followed regardless of their protocol adherence as per the efficacy and safety evaluations:

- Patient withdraws consent or requests discontinuation from the study for any reason
- Termination of the study.
- Lost to follow-up.

Patients who withdraw from this study or are lost to follow-up after signing the informed consent form (ICF) will not be replaced. The reason for patient discontinuation from the study will be recorded on the appropriate case report form.

#### Strategies to improve adherence to interventions

This item is not applicable, since active agent is administered intravenously by health care professionals in a single dose administration.

#### Relevant concomitant care permitted or prohibited during the trial

This study seeks to investigate the effects of CP in addition to standard of care. All concomitant care and interventions are permitted other than concomitant administration of any other experimental treatment.

#### Provisions for post-trial care

No special arrangements for post-trial care are anticipated.

### Outcomes

#### Primary outcome measure

The primary outcome measure is the proportion of patients that progress to categories 5, 6 or 7 (hospitalized severe disease or death categories on the 7-point ordinal scale recommended in the Master Protocol of the WHO R&D Blueprint expert group, at day 15 (Table 1).

**Table 1.** Ordinal scale for illness severity.

|  |
| --- |
| 1. Not hospitalized, no limitation of activities |
| 2. Not hospitalized, limitation of activities |
| 3. Hospitalized, not requiring supplemental oxygen |
| 4. Hospitalized, requiring supplemental oxygen by mask or nasal prongs |
| 5. Hospitalized, non-invasive ventilation or high flow oxygen |
| 6. Hospitalized, intubation and mechanical ventilation or ECMO |
| 7. Death |

#### Secondary outcome measures

The secondary outcome measures include:

- Time to achieve categories 5, 6 or 7 in the 7-points ordinal scale in the 29 days of follow up.
- Time to an improvement of one category from admission using an ordinal scale up to day 29.
- Subject’s clinical status on an ordinal scale at baseline to days 3, 5, 8, 11, 15, and 29.
- Mean changes in the ordinal scale from baseline to days 3,5,8,11,15 and 29.
- Oxygenation free days in the first 28 days (to day 29).
- Ventilator free days in the first 28 days (to day 29).
- Incidence and duration of new mechanical ventilation use.
- Duration of hospitalization (days).
- Mortality rate (all cause) at day 15
- Mortality rate (all cause) at day 29
- Serum level of CRP, lymphocyte count, LDH, D Dimer, IL-6, coagulation tests at baseline and at days 3, 5, 8, 11, 15 and 29.
- Safety assessments of CP+SOC as compared to SOC alone through day 29 considering cumulative incidence of serious adverse events (SAEs), cumulative incidence of Grade 3 and 4 adverse events (AEs), and infusion-related adverse reactions.

#### Exploratory outcomes include the following virology and immunology assessments

a. Qualitative and quantitative RT-PCR for SARS-CoV-2 in nasopharyngeal/oropharyngeal swabs on Days 3, 5, 8, 11 (while hospitalized); and Day 15 and 29 (if able to return to clinic or still hospitalized);
b. Qualitative and quantitative RT-PCR for SARS-CoV-2 in blood on Days 3, 5, 8, 11 (while hospitalized);
c. Quantitative anti SARS-CoV-2 antibodies level at baseline and on days 3, 5, 8, 11 (while hospitalized), and days 15 and 29 (if able to return to clinic or still hospitalized);
d. Neutralizing antibodies study in plasma donors and a subset of patients.

### Participant timelines

The scheduled of interventions and visits can be found in Appendix 2.

### Sample size

We propose an open-label, standard of care controlled, randomised (1:1 ratio); clinical trial with stopping boundaries for efficacy and futility at 20%, 40%, 60 and 80% of the final sample size. The statistical design including the sample size and stopping have been calculated using the East validated software v6.5 by Cytel Inc. The stopping boundaries for superiority and inferiority have been calculated using the Rho family Spending functions.

The primary outcome measure is the proportion of patients that progress to categories 5, 6 or 7 (hospitalized with severe disease or death categories of the 7-point ordinal scale), at day 15. With this design, 278 patients (139 per arm) will be required assuming 20% rate in the control group and an absolute reduction of 10% (10% rate in experimental group), with 80% statistical power a 2.5% one-sided alpha level (5% two-sided). Likewise, approximately 140-200 CP donors will be needed.

With regards to the final sample size, it is predefined that a sample size recalculation will be put in place when 60% of the patients with assessed events at day 15 are available and the 3rd analyses are conducted.

### Recruitment

Patients with COVID-19 will be recruited at the participating clinical sites.

### Assignment of interventions: allocations

#### Sequence generation

Randomization among the two arms will be 1:1 and will be stratified per center.

The randomization process has been developed using the RERAND system integrated within the RDC Onsite ECRF system based on Oracle.

#### Concealment mechanism

This open-label trial will use blind randomization of patients in a 1:1 ratio to CP (+SOC) or SOC alone through a centralized system embedded in the eCRF (Oracle Clinical). Baseline clinical data will be entered in the eCRF before the patient can be randomly assigned via the eCRF at 1:1 ratio to receive standard of care with or without CP as add-on therapy. The system will automatically notify the assigned treatment arm at the eCRF screen and will send a confirmatory message at the email with the randomization information.

#### Implementation

Patients in the treatment arm will receive a single unit of CP (250-300 mL) after randomization on day 1 under control of the patient’s clinician and the Transfusion Service’s hematologist of the hospital. Pathogen reduced plasma bags will be frozen below -25°C, and stored and shipped following European guidelines for blood components storage (36 months below -25°C or 3 months below -18°C). For transport, plasma bags must be kept frozen. No special containers are needed if plasma units are kept frozen at the delivery.

### Assignment of interventions: blinding

This is an open-label study.

### Data collection and management

#### Plans for assessment and collections of outcomes

Investigators are responsible for assessment and collection of outcome, baseline, and other trial data. Data will be entered in the CRF by delegated team members and will be monitored by the clinical research associated. Subjects will be assessed daily while hospitalized. Patients discharged before the end of follow up will be regularly phoned or asked to attend study visits. NP/OP swabs for virus analysis and blood samples for serological analysis will be sent to a central laboratory, where test will be performed according to laboratory standard operating procedures.

#### Plans to promote participants retention and complete follow-up

Patients are free to withdraw from participation in the study at any time. The decision must be communicated and reviewed by investigators. Staff at study sites should explain to these subjects the importance of staying in the study for the full duration of follow-up of this trial for safety reasons. The reason for patient discontinuation from the study will be recorded on the appropriate case report form.

In cases where that a patient becomes lost to follow-up, attempts to contact the patient should be made and documented in the patient’s medical records. Patients who withdraw from this study or are lost to follow-up after signing the informed consent form (ICF) and receiving the study product, will not be replaced.

#### Data management

Data will be examined for compliance with the trial protocol by the monitor and the data manager. Deviations will be sent to the project statistician to plan listings for the Data Review (DR). The objective is to carry out the population selection and definition of the final study populations as well as a preliminary assessment of the quality of the trial data.

All data will be recorded using the defined CRF guidelines for this trial. Also, the blood bank software in each hospital Transfusion Service will be used for recording the plasma units transfused.

#### Confidentiality

Subject confidentiality is strictly held in trust by the participating investigators, their staff, and the sponsor and their agents. This confidentiality is extended to cover clinical information relating to subjects, test results of biological samples and all other information generated during participation in the study. All study data and research specimens that leave the site (including any electronic transmission of data) will be identified only by a coded number that is linked to a subject through a code key maintained at the clinical site. All source records including electronic data will be stored in secured systems.

No identifiable information concerning subjects in the study will be released to any unauthorized third party. Subject confidentiality will be maintained when study results are published or discussed in conferences. The study monitor, other authorized representatives of the sponsor, representatives of the Research Ethics Committees (RECs), and/or regulatory agencies may inspect all documents and records required to be maintained by the investigator, including but not limited to, medical records (office, clinic, or hospital) for the subjects in this study. The clinical study site will permit access to such records.

#### Plans for collection, laboratory evaluation, and storage of biological specimens for genetic or molecular analysis in this trial/future use

Samples will be collected by investigators or designees. After that, handling, labeling, processing, storaging and/or shipping according to protocol will be performed and the samples will ultimately be sent to the central laboratory.

The Sponsor and the center, may use data and samples for future research projects related with COVID-19, taking the appropriate measures to ensure the protection of their privacy and will not allow their encrypted data to cross with other data bases that could allow their identification. Any future studies will be previously evaluated by a Research Ethics Committee and will comply with the applicable ethical and legal requirements.

### Statistical methods

#### Statistical methods for primary and secondary outcomes

A detailed Statistical Analysis Plan (SAP) agreed upon by the CT Executive Board and the Project Statistician will be available early during the recruitment phase. This SAP will follow the general regulatory recommendations given in the ICHE9 (CPMP/ICH /363/96) guidance, as well as other specific guidance on methodological and statistical issues [7].

Likewise, it will stick to the recommendations given by the consensus documents of the scientific journals to improve reliability and value of medical research literature by promoting transparent and accurate reporting of clinical research studies.

The proportion of patients with failure, defined as categories 5, 6 or 7 of the 7-point ordinal scale at day 15, will be estimated using a log-binomial regression model including stratification variables. In the unexpected event that the model does not fit, the Poisson regression model with long-link and robust variance estimator will be used instead.

Binary efficacy and safety outcomes will be analysed as described for the primary endpoint.

Kaplan-Meier model will be used to analyse survival endpoints (TTF and OS). In all these analyses, in addition to the Kaplan-Meier curve, median, Q1, Q3 and their corresponding 95% CI, number of events and censored cases distribution will be shown. Group comparisons will be done using the (stratified) log-rank test and the (stratified) hazard ratios -HR- (95%CI) from the Cox model.

#### Interim analyses

Interim analyses for comprehensive efficacy (or futility) and safety data monitoring analyses will be conducted when 20%, 40%, 60% and 80% of patients have been recruited and completed day 15 assessment, or at the discretionary criteria of the DSMB, when needed. The study may be stopped prematurely if either the efficacy or the futility boundaries are crossed. The boundaries will be adapted to the actual information rates in each inspection using the Rho family Spending functions with (rho=7) implemented in the East validated software v6.5 (or later releases) by Cytel Inc. up.

Ad hoc reviews will be undertaken at any time if there are other specific safety concerns. The study will not stop enrolment awaiting these DSMB reviews, though the DSMB may recommend temporary or permanent cessation of enrolment based on their safety reviews.

#### Methods for additional analyses (e.g. subgroups)

Subgroup analyses will be performed to assess the impact on efficacy of two different key factors, i.e. the level of neutralizing antibodies in the administered plasma and the timing of the disease i.e. early or late stages considering 7 days as cut-off, viral load and IgG or IgM. The following strategy will be conducted before splitting the analysis into subgroups: test of the overall treatment effect, test of the treatment-by-subgroup interaction at the 10% level of significance or test of the treatment effect in each subgroup category

#### Methods to handle missing data

In principle, the rate of missing data is estimated to be very low due to the type of endpoint, easily available with a fast-clinical assessment, so no impact on the primary analysis is expected. In any case, a very conservative strategy will be implemented consisting of imputing any missing data or other binary efficacy secondary outcomes will be considered to failures, irrespectively to the reason for missingness. With regards to the continuous variables, mixed models ([8]–[11]) are robust to the presence of missing at random (MAR) and conducts the analysis with all participants despite the presence of missingness. Of note, this method calculates the estimations based on the variance-covariance structure but without any formal imputations.

#### Plans to give access to the full protocol, participant-level data, and statistical code

These plans are not yet in place.

### Oversight and monitoring

*Composition of the coordinating centre and trial steering committee*

▪ Coordinating centre The study is led by the Hospital Universitario Puerta de Hierro Majadahonda.
▪ Trial steering committee

The trial steering committee consists of the following members:

- Dr. Rafael Duarte, Hematology and Hemotherapy Department. Hospital Universitario Puerta de Hierro Majadahonda
- Dr. Cristina Avendaño Solà, Clinical Pharmacology Department. Hospital Universitario Puerta de Hierro Majadahonda
- Dr. Antonio Ramos-Martínez, Internal Medicine Department. Infectious diseases Unit. Hospital Universitario Puerta de Hierro Majadahonda
- Dr. José Luis Bueno, Hemotherapy & Apheresis Units. Hematology and Hemotherapy Department. Hospital Universitario Puerta de Hierro Majadahonda
- Dr. Elena Múñez, Internal Medicine Department. Infectious diseases unit. Hospital Universitario Puerta de Hierro Majadahonda
- Dr. Belén Ruiz-Antorán, Clinical Pharmacology Department. Hospital Universitario Puerta de Hierro Majadahonda
- Dr. Rosa Malo de Molina, Servicio de Pneumology. Hospital Universitario Puerta de Hierro Majadahonda
- Dr. Ferrán Torres, Clinical Pharmacology Department. Hospital Clínic Barcelona. Medical Statistics core facility - IDIBAPS.
- Dr. Inmaculada Casas Flecha, Flu and Respiratory Virus Unit. Centro Nacional de Microbiología, Instituto de Salud Carlos III

#### Trial monitoring

The Spanish Clinical Research Network (SCReN) is responsible for project management, regulatory compliance and trial monitoring.

#### Data management team

PIVOTAL is the CRO responsible for data management, preparation of the eCRD, quality assurance, and preparation of the SAP.

### Composition of the data monitoring committee, its role and reporting structure

The independent DSMB in this study is responsible for reviewing the reports regarding the safety and efficacy of the study patients protocol adherence and making recommendations to continue or terminate the study or to modify the sample size of the basis of the results from the interim analysis. The DSMB members are all independent of the sponsor and have no financial or other conflict of interest.

### Adverse event reporting

Serious adverse events (SAEs) and grade 3 or 4 adverse events will be collected from the time of informed consent to day 29. SAEs will be followed up until the SAE has subsided, returned to baseline, or is stable. Infusion-related adverse reactions will be recorded within 24 hours after the end of plasma administration by a trained Hemovigilance nurse or physician, according to the Active 24h quarantine Hemovigilance Program (HEMACUA)

Investigators will be instructed to actively monitor the occurrence of prespecified adverse events os special interest: TRALI (Transfusion-related acute lung injury), ADE (Antibody-dependent enhancement of infection and TACO (Transfusion-associated cardiac overload).

### Plans for auditing trial conduct

Monitoring for this study will be performed by the sponsor and SCReN. Monitoring online visits will include, but not limited to, review of regulatory files, accountability records, CRFs, ICFs, medical and laboratory reports, site study intervention storage records, training records, and protocol and GCP compliance.

On site and,off-site monitoring, central review of data collection and remote source data verification will be allowed according to EMA and AEMPS guidance/guidelines on the management of clinical trials during the covid-19 (coronavirus) pandemic.

### Plans for communicating important protocol amendments to relevant parties (e.g. trial participants, ethical committees)

During the trial, any amendments to the protocol or consent materials will be approved by the REC before they are implemented.

### Dissemination plans

Following completion of the study, the results will be published in a scientific journal. Nevertheless, due to the critical need of results during the current epidemic COVID-19 crisis, preliminary results will be released by the sponsor to the Health Authorities.

### Ethical and regulatory

The clinical study will be conducted in accordance with the relevant national and international good clinical practice (GCP) guidelines, and the Declaration of Helsinki, each in the applicable version. The study protocol and the donors and the patients ‘written informed consent were submitted to and approved by the Research Ethics Committee of Hospital Puerta de Hierro Majadahonda on March 23th, 2020.

## Discussion

COVID-19 is a respiratory disease caused by a novel coronavirus (SARS-CoV-2) and causes substantial morbidity and mortality. At the time the study protocol was designed, there were no vaccines to prevent COVID-19 or infection with SARS-CoV-2 or therapeutic agent to treat COVID-19. Indeed, as in any other health emergency, there was an enormous pressure to find cures and stop COVID-19 epidemic. This has prompted the extensive use of unproven treatments either on a compassionate use basis, in observational studies, or even in small clinical trials, but these do not provide the requires level of evidence to solve the existing uncertainties. Thus, there was a substantial need to conduct appropriately powered randomized controlled studies in order to generate a reliable and conclusive evidence regarding the benefits and risks of these therapies.

In this scenario, convalescent plasma was hypothesized to be a potential therapeutic option given its extensively recognized immunomodulatory and anti-inflammatory effects. However, the actual benefits and risks of the intervention remain to be established, particularly in this novel condition. Currently, there are over 250 studies registered in ClinicalTrials.gov about convalescent plasma as a treatment for respiratory disease caused by COVID-19 all over the world, most of which are non-controlled clinical studies, that are ongoing. Therefore, in accordance with the WHO Blood Regulators Network position statement [12] and our commitment to generate compelling evidence, we planned to conduct the proposed randomized and controlled trial to establish the effectiveness and safety of convalescent plasma from disease survivors in the treatment of patients with severe COVID-19 pneumonia.

This clinical trial is designed to evaluate passive immunotherapy with convalescent plasma for the treatment of adult patients hospitalized with non-severe COVID-19. With regard to the regulatory aspect, convalescent plasma from a single donor is not considered a medicinal product but rather it is subject to the regulation applicable to blood transfusions at the EU level. Subject to demonstration of its efficacy, it was considered that CP would constitute a universally accessible treatment option given that it relies entirely on already existing transfusions systems and technical requirements that are already established in every country. This makes CP a particularly attractive potential therapeutic option; even more considering that it is possible that the results of this study will contribute to the establishment of clinical recommendations to treat similar conditions.

The study was designed as an open–label study due to ethical implications of a sham transfusion in the context of COVID-19 pandemic. The study was also designed as a controlled clinical trial. Initially, a three-treatment arm study including no experimental treatment (only SOC), unspecific standard plasma (plus SOC), and CP(+SOC) was considered. The aim was to demonstrate that any potential benefit from CP treatment was due to the presence neutralizing antibodies and any other related cytokines released in a successful immune response and not just to the existing non-specific immune-components in SARS-CoV-2 non-exposed donors. The inclusion of a third treatment arm with unspecific plasma might have also allowed ruling out any potential deleterious effect of transfusing an enriched plasma to a subject developing his/her own immune response. Nevertheless, a three-treatment arms trial was deemed unrealistic due to practical reasons and also to the well-known adverse events related to blood component transfusion; some of them even fatal; specially transfusion related acute lung injury (TRALI).

Concerning the study population, a potential role of immunotherapy across the spectrum of COVID-19 disease, has been suggested and several studies are ongoing to test different hypothesis. When defining the study population for our study, we considered that the best candidates for passive immunotherapy would be patients hospitalized due to COVID-19 pneumonia, thus in need for treatment, but early in the course of the disease when a primary immune response is not yet established. As a result, only patients requiring hospitalization for COVID-9 pneumonia without mechanical ventilation (invasive or non-invasive) or high flow oxygen devices, and with no more than 12 days between the onset of symptoms (fever or cough) and treatment administration day were allowed to enroll. It was believed that this early intervention would allow patients to benefit from the immune response of a subject who had developed a successful one, which might help clearing SARS-CoV-2 virus and to prevent progression to a more severe condition. On the other hand, patients requiring ventilation (mechanical or non-mechanical) were excluded given that they are less likely to respond to passive immunotherapy and more likely to develop ADE (antibody –dependent enhancement of disease), a worrisome complication seen previously in other viral infection (13) where a high specific antibody infusion could trigger a severe life-threatening immune response.

With regard to convalescent plasma donor’s selection, processing and storage, it has been performed following the European and Spanish guidelines (RD 1088/2005) for standard plasma donation; including a pathogen reduction (PR) treatment. Lack of process control and standardization has been raised as one of the reasons why clinical trials have failed so far. We will select CP with anti SARS-CoV-2 IgG antibodies using a validated Enzyme Linked Immunoabsorbent Essay (ELISA); with an IgG amount above a standardized cut-off. These assays have been performed at the trial central laboratory. No previous determination for neutralizing antibodies titers will be used to select CP in our trial, as this would be the norm in the use of CP in the midst of a pandemic. Titration of neutralizing antibodies in the administered plasma will be subsequently performed and its relation to the outcomes could provide useful information about efficacy and safety of CP.

An important issue in our trial is that we selected our CP donors from recovered mild COVID19 conditions. Although patients recovered from severe COVID19 was not an exclusion criteria for donors in our trial, we chose mild donors based in two assumptions: first, severe patients could have a high anti SARS-CoV-2 IgG titer, but low neutralizing antibody titers; second, high neutralizing antibodies titers in CP could increase the risk of ADE (antibody–dependent enhancement of disease). Thus, our donor’s selection strategy is aimed to select CP with medium levels of antibody titers, which is thought to provide a fair balance of efficacy and safety. CP will be administered as an add-on to the standard of care as defined in each study center. Based on the clinical practice recommendations in place at the time the study was designed, the standard of care could be based on any of the followings: lopinavir/ritonavir, darunavir/cobicistat, hydroxy/chloroquine, tocilizumab. Remdesivir was added to standard of care once it was available outside clinical trials. Demonstration of efficacy in this context is particularly challenging, but ethical and feasibility issues were prioritized.

Defining the primary endpoint posed additional challenges given the existing uncertainties around the natural course of the disease, and the need to balance hard clinically relevant outcomes against outcomes that might occur earlier. The ordinal scale recommended by the WHO for clinical trials in COVID-19 was finally selected, as it was considered to provide a reasonable balance between these two criteria, while facilitating any external comparison with similar clinical trials outcomes. We also include a range of secondary endpoints to assess the scope of the disease and the potential benefits/risks of treatment in a broader spectrum, in an attempt to show consistency and the robustness of any potential effect shown in the primary endpoint.

Finally, to overcome practical difficulties due to paper handling limitations at COVID wards but to still comply with ethical requirements applied to clinical research, oral witnessed consent will be accepted before entering the trial. Written consent form will be obtained from the patient himself or acceptable representatives as soon as possible.

### Trial status

ConPlas-19 Protocol version 2.2 as of 22nd April 2020. Patient’s recruitment started on 4th April 2020, donor’s recruitment started on 2nd April 2020. As of 10th July, 2020 patients’ recruitment has been temporarily interrupted awaiting some study modifications. Patients already recruited continue under evaluation. It is anticipated that recruitment will be complete by end of 2020.

## Data Availability

The datasets generated and/or analyzed during the current study will be made available. The corresponding authors will evaluate any request for data sharing and will consult with the steering committee after the publication of the main results.

## Declarations

### Ethics approval and consent to participate

The study protocol and the donors and the patients’ informed consents were submitted to and approved by the Research Ethics Committee of Hospital Puerta de Hierro Majadahonda on March 23th, 2020 (REC number PI 57-20)

I herewith certify that this trial has received ethical approval from the appropriate ethical committee as described above. Consent from participants to participate in the study will be obtained before any study procedure.

### Consent for publication

Not applicable.

### Availability of data and materials

The datasets generated and/or analyzed during the current study will be made available. The corresponding authors will evaluate any request for data sharing and will consult with the steering committee after the publication of the main results. Requests can be sent to.

## Authors’ contributions

CAS, RDP, JLB, BRA, FT, EM, ARM, AFC, IC, CPH did the literature search, conceived the study design. CAS acted as the study sponsor and principal investigator. RDP acted as principal investigator. ARM also acted as clinical trial national coordinator. JLB acted as plasma production coordinator. AVI also acted as project manager. ISD was crucial for trial organization and execution and revised the manuscript. EDS, ASL, and BRA drafted the manuscript, made the tables and figures, and had final approval of the manuscript. All authors critically revised the manuscript

## Acknowledgements

Not applicable

## Authors’ information

Not applicable

## Conclusion

Due to the existing uncertainties on the potential role of convalescent plasma in adult patients with severe COVID-19 pneumonia, we consider that the publication of the study protocol will help other researchers to understand the rationale behind our clinical trial design and may contribute to define future research strategies with CP in the field of COVID-19 or other viral diseases.

